# Genetic Semantic Dementia? Twins’ Data and Review of Autosomal Dominant Cases

**DOI:** 10.1101/2024.10.18.24313757

**Authors:** Shalom K. Henderson, Matthew A. Lambon Ralph, P Simon Jones, Michelle Naessens, Thomas E. Cope, David J. Whiteside, Arabella Bouzigues, Lucy L. Russell, Phoebe H. Foster, Eve Ferry-Bolder, John van Swieten, Lize Jiskoot, Harro Seelaar, Raquel Sanchez-Valle, Daniela Galimberti, Matthis Synofzik, Barbara Borroni, Jonathan D. Rohrer, GENetic Frontotemporal dementia Initiative (GENFI), James B. Rowe, Karalyn E. Patterson

## Abstract

**Background:** Amongst different subtypes of frontotemporal dementia (FTD), semantic dementia (SD, also known as the semantic variant of primary progressive aphasia, svPPA), is the least likely to have a genetic basis.

**Methods:** Our study had two aims: (i) to describe two SD cases and detailed assessments of their unaffected monozygotic (MZ) twins, and (ii) to review cases with FTD-associated mutations or known family history classified as SD/svPPA either in the Genetic Frontotemporal dementia Initiative (GENFI) or in the published literature.

**Results:** The two affected twins displayed characteristic features of SD, both in neuroimaging and cognition, whereas their MZ twins exhibited no abnormalities in either regard, even up to 15 years of follow-up for one affected twin. Only five cases out of more than 1300 people in GENFI were classified as svPPA, with a genetic mutation. The systematic review revealed 29 cases with sufficient clinical and language details regarding ‘genetic’ SD/svPPA. A comparison of these five GENFI and 29 literature cases to the patterns observed in a large number of sporadic cases revealed critical differences in presentation.

**Conclusions:** Both parts of our study suggest that true SD/svPPA is unlikely to have an autosomal dominant genetic aetiology and that, while mutation carriers may resemble SD/svPPA in some respects, they may not meet current clinical diagnostic criteria for this condition.

**What is already known on this topic**: Approximately 30% of all frontotemporal dementia cases are associated with an autosomal dominant pattern of inheritance but the reported prevalence of mutation in semantic dementia/semantic variant of primary progressive aphasia (SD/svPPA) is very low.

**What this study adds**: From (a) outlining the discordance for SD/svPPA in two pairs of monozygotic twins and (b) comparing the clinical and cognitive profiles of people who had been classified as SD/svPPA with an FTD-associated genetic mutation *versus* sporadic SD/svPPA, we conclude that true SD/svPPA is unlikely to have an autosomal dominant genetic aetiology.

**How this study might affect research, practice or policy**: Our study highlights gaps in the understanding of environmental and epigenetic influences on sporadic SD/svPPA and a need for further unbiased genotyping and phenotyping of SD/svPPA and “SD-like” syndromes.

**Open access:** For the purpose of open access, the authors have applied a CC BY public copyright licence to any Author Accepted Manuscript version arising from this submission.

## Introduction

Genetic mutations are important causes of many types of frontotemporal dementia (FTD), with approximately 30% of cases described as having an autosomal dominant pattern of inheritance.^1–3^ Genetic FTD typically has a mutation in one of three genes: (1) microtubule-associated protein tau (MAPT), (2) progranulin (GRN), or (3) the chromosome 9 open reading frame 72 (C9orf72).^4^ The behavioural variant frontotemporal dementia (bvFTD) is more likely to be due to a mutation than other FTD syndromes.^5^ The reported prevalence of mutations is relatively low in primary progressive aphasia (PPA), and especially low in semantic dementia (SD) or the semantic variant of PPA (svPPA).^6^ A suggestive family history is identified in only around 2–7% of SD/svPPA cases.^3,7^

This paper has two complementary aims: first, to report detailed evaluation of two new SD cases and their monozygotic (MZ) twins; second, to review the clinical features of people described as SD/svPPA due to an FTD-related mutation and with a definite or possible family history, reported in either the Genetic Frontotemporal dementia Initiative (GENFI) or the published literature.

Twin studies serve as a window into the manifestations of autosomal dominant inherited dementia. On the one hand, similar clinical and neuroimaging findings have been reported in MZ twin pairs. For example, McDade and colleagues found concordant clinical, neuroanatomical, and serum progranulin levels in a pair of MZ twins with GRN mutations.^8^ Even amongst MZ twins, however, a diagnosis of Alzheimer’s disease, Parkinson’s disease, or dementia with Lewy Bodies in one twin does not reliably predict the same diagnosis in the other.^9–11^ C9orf72- and MAPT-associated PPA are rare.^12^ GRN mutations are the most common genetic cause of PPA, although usually presenting with non-fluent agrammatic or mixed features rather than the clear syndrome of SD/svPPA.^13^ Mutations have been reported in a small percentage of people with seemingly sporadic PPA, including SD/svPPA^14^ and non-fluent variant PPA,^15^ but their clinical phenotypes might not be typical of classical (non-genetic) forms of PPA. Although clinico-pathological correlation is very high for sporadic svPPA, clinical-genetic relationships may be different. Accordingly, in this systematic review we consider whether the patients with a mutation/family history who have been described as SD/svPPA differ in notable ways from the paradigmatic sporadic phenotype.

### Part 1. Case reports of MZ twins

Twin data were acquired under the PrEPPAReD protocol, approved by the Cambridge 2 Research Ethics Committee (REC reference: 07/Q0102/3; Title: Prospective Evaluation of Parkinson Plus & Related Disorders, formerly entitled: Diagnosis and prognosis in Progressive Supranuclear Palsy (PSP), Corticobasal degeneration (CBD) and Dementia). The affected twins had provided consent to research before losing mental capacity and the decision to include them in research after the loss of mental capacity was affirmed by their personal consultee as set out in the Health and Social Care Framework law for England and Wales. All healthy participants provided written informed consent.

<u>Case A</u>: An individual in the early 60s presented to a Neurology Clinic with difficulty in naming objects and people. A was especially troubled by the inability to remember the names associated with a favourite hobby. A’s past medical history was unremarkable apart from mild hypertension. A had a monozygotic (MZ) twin sibling who was reported to be healthy. There was no report of cognitive problems in either of their parents. A Consultant Neurologist made a preliminary diagnosis of SD and referred A to a specialist centre in FTD. Assessment confirmed severe anomia: 10/64 on the Cambridge Semantic Battery (CSB) naming test with poor scores on both the living (5/32) and manmade (5/32) objects. On the more difficult Graded Naming Test, A could not name any of the 30 pictures. A’s semantic impairment was severe in receptive as well as expressive tasks: on the easy word-to-picture matching task in the CSB, A’s score was 28/64. The Repeat-and-Point Test requests the responder first to repeat a spoken multisyllabic target word (e.g., *rhinoceros*, *stethoscope*) and then to point to the corresponding picture in an array of seven pictures from the same semantic category. A correctly repeated 9/10 of the target words aloud (making a minor phonological error on *rhododendron*) but only pointed correctly to 4/10 pictures. A’s Mini Mental State Examination (MMSE) was 24/30 and, on the Addenbrooke’s Cognitive Examination – Revised (ACE-R), A’s overall score was 52/100, with good orientation, registration, perceptual and visuospatial performance, but very poor scores on any component requiring semantic knowledge or word memory, poor fluencies (category worse than letter) and surface dyslexia (e.g., *sew* read aloud as “sue”). By contrast, non-semantic/language aspects of assessment revealed good performance. A made no errors on the Trail Making Test, with times to complete these within the normal range (Trails A: 37 seconds, Trails B: 86 seconds). A’s digit span was 6 forwards and 4 backwards. A’s Rey Figure copy was good (34/36), but Rey Figure recall was poor and lacking in detail (8/36). A was fully self-caring, reported no difficulty with route finding (confirmed by A’s next of kin) and showed no neuropsychiatric features or behavioural symptoms. An MRI showed typical bilateral but left predominant anterior temporal lobe atrophy (Figure 1).

**Figure 1.**
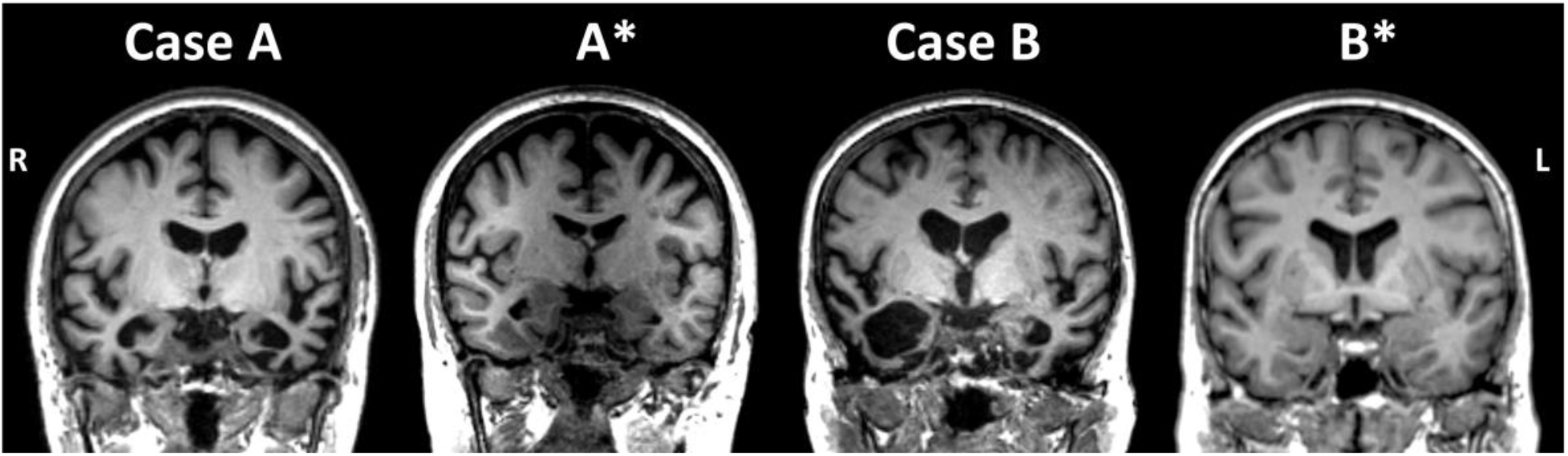
Neuroimaging for the twin pairs. Coronal MRI slices through the anterior temporal lobe. Age range at scan was between 61 and 65 for A, between 71 and 75 for A*, between 66 and 70 for B, and between 66 and 70 for B*. Note that A*’s scan was 12 years after A’s initial scan.

Over the next few years, A’s family began to report mild impairment in A’s housework and hobbies/interests, fidgeting and loss of insight. After 2 years, A’s MMSE and ACE-R were 20/30 and 41/100, respectively. By three years, anxiety and agitation were appearing, and A was developing fixed routines and clock-watching and beginning to hoard some items; but A was safe at home alone, remembered to take regular medication (anti-hypertensive) and, although now retired, was still doing basic chores at home. Despite the addition of some behavioural features with progression, the salient feature of both A’s initial presentation and the pattern of decline was profound semantic loss. By 4 years after presentation, A’s MMSE and ACE-R were 7/30 and 17/100, respectively. A was seen regularly at the specialist centre in FTD until the COVID pandemic, by which time A was nonverbal and dependent for activities of daily living (ADLs).

#### A*, MZ twin of Case A

Thirteen years after A’s presentation, and based on a report from A’s family that A’s twin A* remained completely healthy, A* was invited to undergo cognitive testing and structural MRI at the specialist centre in FTD (see Figure 1). A* scored 29/30 on the MMSE and 89/100 on the ACE-R, with ACE-R subdomains for Language and Memory of 24/26 and 20/26, respectively. A* made no errors on the Trail Making Test, with times of 27 seconds for Trails A and 85 seconds for Trails B. On the Pyramids and Palm Trees semantic association test, A* scored 51/52, and on the difficult Graded Naming test, 23/30, well within the normal range. Digit span was 6 forwards and 4 backwards. Fluency responses for the initial letters of F, A, S were 10, 9, 13 words in 60 seconds each. On examination, A* had no neurological or neuropsychiatric signs or symptoms. MRI was normal (Figure 1).

<u>Case B</u>: An individual in the late 60s was referred by a general practitioner to the specialist centre in FTD, with a 2-year history of deteriorating “memory”. The particular problems noted in the referral were (a) forgetting people’s names and (b) difficulty in following the story lines in TV programmes, with repeated questions of “What’s going on?”. Besides mild anxiety with depression, B had no neurological or psychiatric history. B remained independent in all ADLs, was using a computer without difficulty and was still doing the household accounts. On the other hand, B had become fixated on games on the mobile phone and on jigsaw puzzles; was restless and walking for significant periods of time (never getting lost); and had lost interest in previous activities. B’s initial MMSE was 27/30, and ACE-R score was 73/100, with normal sub-scores for Attention, Orientation and Visuospatial Function, but significant deficits for Memory, Fluency and Language, including surface dyslexia. B named 6/12 pictures of objects and animals. Cranial nerve and limb examination was normal. MRI revealed bilateral anterior temporal atrophy, greater on the right (Figure 1).

After six months B’s MMSE score remained 27/30 but an ACE-R score was 60/100, with the same pattern of preserved Attention, Orientation and Visuospatial skills, but very poor Memory, Fluency and Language; B’s naming of objects and animals had dropped from 6 to 3/12. On drawing-to-name, B’s animal drawings lacked characteristic features (e.g., no elephant-specific attributes such as the trunk, tusks, large ears, or thick legs). B’s face recognition was extremely poor: in a set of 20 photographs of very famous people, B recognised only Queen Elizabeth II and Elvis Presley. At 2 years’ follow-up, B was cheerful and talkative with the characteristically empty speech of SD. B’s comprehension was poor with, for example, failure to know the meaning of words like *understanding, flavour, caterpillar,* or *sparrow*.

#### B*, MZ twin of Case B

In response to the request of the specialist centre, B* expressed willingness to be assessed and underwent cognitive testing and structural MRI soon after B. There was no neurological or psychiatric history. On examination, B* displayed no neurological deficits, and cognitive testing was entirely normal: 96/100 on the ACE-R (with perfect object naming), 51/52 on Pyramids and Palm Trees, digit span 6 forwards and 5 backwards. The family of B* gave no endorsements of cognitive or behavioural abnormalities on the Cambridge Behavioural Inventory. MRI scan was normal.

### Part 2. Review of cases described as SD/svPPA with mutation/family history

#### GENFI patients

Patient data for cases classified as svPPA were drawn from the international Genetic FTD Initiative (GENFI), which consists of 44 research centres in Europe and Canada.^16^ For data acquisition, all GENFI sites had local ethical approval for the study from relevant institutional review board or research ethics committee and all participants provided written informed consent. Analyses of GENFI data were conducted under the Cambridge Genetic Frontotemporal Dementia Initiative protocol approved by the Cambridge 2 Research Ethics Committee (REC reference: 17/EE/0032; title: Cambridge Genetic Frontotemporal Dementia Initiative).

Five symptomatic patients with a genetic mutation were categorised as svPPA^17^ and included in the present review. All participants underwent a standardised clinical assessment including a history, the Mini-Mental State Examination (MMSE), the CDR® plus NACC FTLD global score, and the CDR® plus NACC FTLD sum of boxes which is used to classify mutation carriers as either presymptomatic (global score of 0, asymptomatic, or 0.5, prodromal) or fully symptomatic (score ≥ 1).^12^ Language was assessed by a clinician using the GENFI clinical questionnaire, which includes the Progressive Aphasia Severity Scale (PASS).^18^ More detailed neuropsychological assessment data were available for four of the five patients and structural MRIs for three.

These five genetic patients were compared with a sporadic sample of svPPA patients (*n* = 19) from Samra *et al*.^12^ who had undergone the same standardised GENFI clinical assessment mentioned above. The mean age of the sporadic sample was 64 and diagnosis of PPA, as well as classification of svPPA, were supported by current consensus criteria.

#### Statistical analysis

We employed a Bayesian approach to test whether the performance and profile of the five genetic svPPA cases reported in GENFI were meaningfully different from the sporadic svPPA sample.^12^ Using the point and interval estimates of effect sizes for single case to normative sample design in neuropsychology,^19^ we derived a point estimate of the effect size for the difference between each case and the sporadic sample with an accompanying 95% credible interval, as well as a point and interval estimate of the abnormality of the case’s score (i.e., the percentage of the sporadic sample that would obtain a lower score than the genetic case).

#### Literature search and selection criteria

We searched PubMed for articles on genetic frontotemporal lobar degeneration from database inception up to August 2023 using the following terms: (((c9orf72) OR (chromosome 9 open reading frame 72) OR (grn) OR (granulin) OR (progranulin) OR (MAPT) OR (microtubule associated protein tau))) AND (((semantic dementia) OR (semantic variant primary progressive aphasia) or (SD) or (svPPA))) AND ((mutation) OR (expansion)). From the resulting 2,684 papers, we screened for studies including one or more SD/svPPA cases. Next, 249 full-text articles were assessed for eligibility based on (1) inclusion of genetic cases with SD or svPPA and (2) sufficient clinical and/or language details available. Focussing on cases with SD^20^ or svPPA,^17^ twenty publications were included in this review.

Our systematic review table includes details about: the type of mutation; age at onset, first visit and death; family history; handedness; country of residence; first reported symptom; presence of isolated semantic deficits; additional language, cognitive and behavioural deficits and/or other notable symptoms; imaging findings, including atrophy in the anterior temporal lobes; and pathology if reported. We also indicated whether the patient *presented* with an isolated semantic deficit, if this information was available, and coded the response for specific cognitive-behavioural deficits as “initial” and “later”.

For family history, we calculated the modified Goldman score^3,6,21^ where a minimum score of 1 indicates a strong autosomal dominant family history of FTD, motor neuron disease, corticobasal syndrome or progressive supranuclear palsy, with at least three affected people in two generations with one person being a first-degree relative of the other two. A maximum Goldman score = 4 denotes no or unknown family history.

## Results

### GENFI

As an initial comment, it is noteworthy that only five people out of more than 1300 in GENFI were classified as svPPA.

#### Demographics

Demographic details for the five cases are shown in Table 1. The GENFI family history questionnaire includes diagnostic questions about the participant’s parents, siblings, and children. One of Case 1’s parents had a diagnosis of dementia not otherwise specified (NOS) with age of onset in the mid 70s and age of death in the mid 80s. Out of three siblings, only one was affected with a diagnosis of bvFTD with age of onset in the mid 50s. One of Case 2’s parents had a diagnosis of dementia NOS with a late age of onset. One of Case 5’s parents had a diagnosis of dementia NOS with a young age of onset and a middle age of death. All three siblings were affected but their diagnoses were indicated as “other” with no details for age of onset or death. Further details from the GENFI clinical report can be found in Supplementary Table 1 and highlight notable differences between genetic and sporadic cases: (1) genetic cases exhibited unexpected impairments such as deficits in articulation (i.e., Case 4’s PASS articulation score), reduced fluency in speech (i.e., Cases 1 and 4’s PASS fluency scores), grammar/syntax (Cases 1, 2, and 4’s PASS grammar/syntax scores), and repetition (Cases 1, 2, and 4’s PASS repetition scores); and (2) some of the typical or expected impairments in sporadic SD were missing in the genetic cases; for example, Cases 2, 3 and 5 apparently had preserved sentence comprehension (Cases 2, 3, and 5’s PASS sentence comprehension scores) and no reading impairment (i.e., Cases 1, 3, 4, and 5’s PASS dyslexia scores). Figure 2 shows the structural MRIs for Cases 2, 3 and 5 at their initial assessment, with cases 3 and 5 lacking the temporal lobe asymmetry that is often seen in SD.

**Figure 2.**
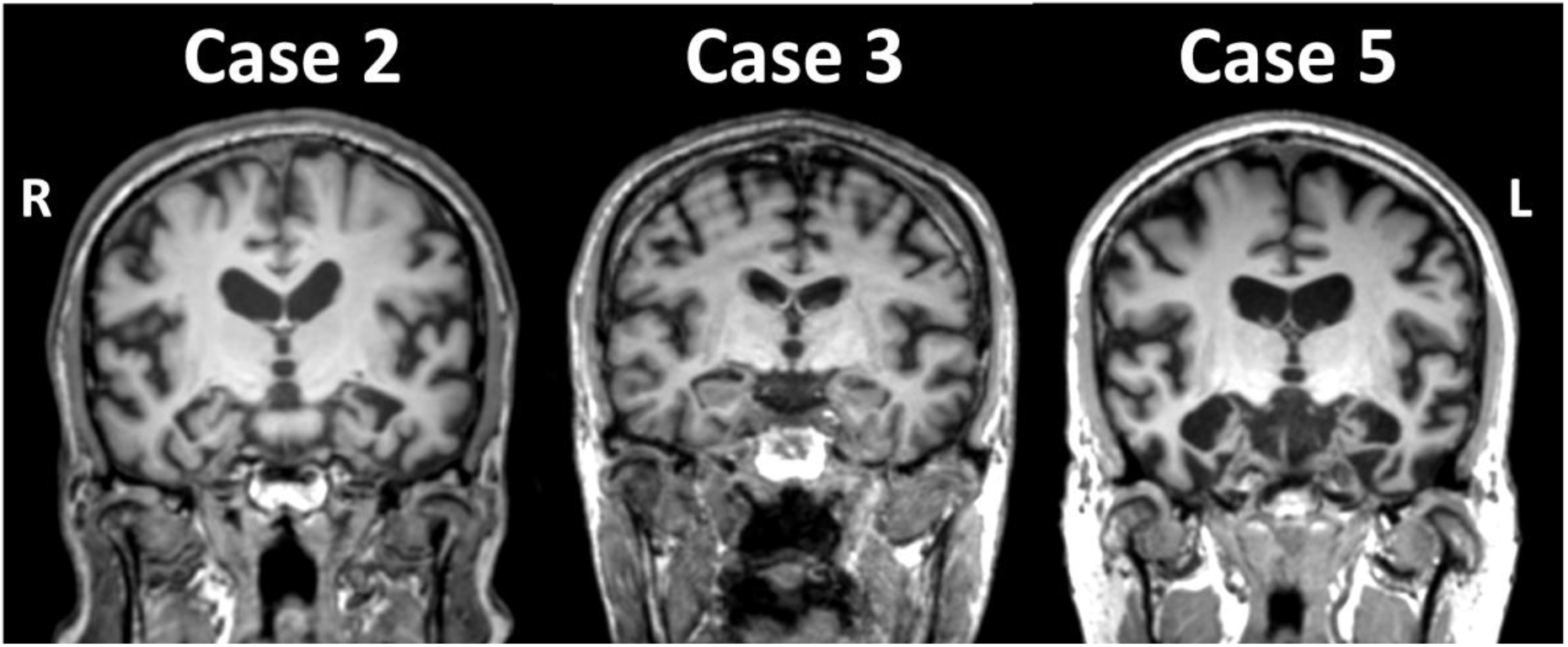
**Coronal T1-weighted MRI of GENFI cases at their initial assessment**.

**Table 1.** Demographics for the GENFI svPPA cases recorded at the initial study session

| Case | Genetic Group | Mutation | Handedness | Ethnicity | Education (years) | Age at onset in ranges | Age at first visit in ranges | Family history | Imaging |
| --- | --- | --- | --- | --- | --- | --- | --- | --- | --- |
| 1 | C9orf72 | NA | R | White | < 10 | 51 - 55 | 56 - 60 | Yes | No |
| 2 | C9orf72 | NA | R | White | ≥ 10 | 71 - 75 | 71 - 75 | Yes | Yes |
| 3 | C9orf72 | NA | R | White | < 10 | 56 - 60 | 56 - 60 | No | Yes |
| 4 | GRN | C253X | R | White | ≥ 10 | 61 - 65 | 66 - 70 | No | No |
| 5 | MAPT | P397S | R | White | ≥ 10 | 56 - 60 | 56 - 60 | Yes | Yes |

#### Neuropsychological assessment, clinical signs and language

The Bayesian comparisons between each genetic case and the sporadic svPPA sample from Samra and colleagues^12^ are shown in Table 2. Some test scores were not available due to assessment differences between GENFI 1 and GENFI 2. Cases 1 and 2 participated in GENFI 1 and cases 3 to 5 in GENFI 2. Case 4 exhibited a significantly high PASS sum of boxes score and a strikingly low score on the MMSE compared to the sporadic svPPA sample. The scoring for PASS ranges from 0 (normal) to 3 (severe impairment). The two-tailed probability of 0.002 for PASS sum of boxes indicates that the patient’s score was significantly (*P* < 0.05) below the sporadic mean and that an estimated 0.2% or less of the sporadic SD sample would obtain a score lower than patient 4. This is also indicated in the Bayesian estimated percentage of 99.9%; in other words, nearly 100% of cases in the sporadic sample are expected to have a lower PASS sum of boxes scores, which for this measure indicates *less* overall language impairment. By contrast, Case 4’s MMSE had a two-tailed probability of 0.01 and a Bayesian estimated percentage of 0.05%: in other words, the majority of sporadic cases would be expected to have a higher MMSE score than Case 4.

**Table 2.**
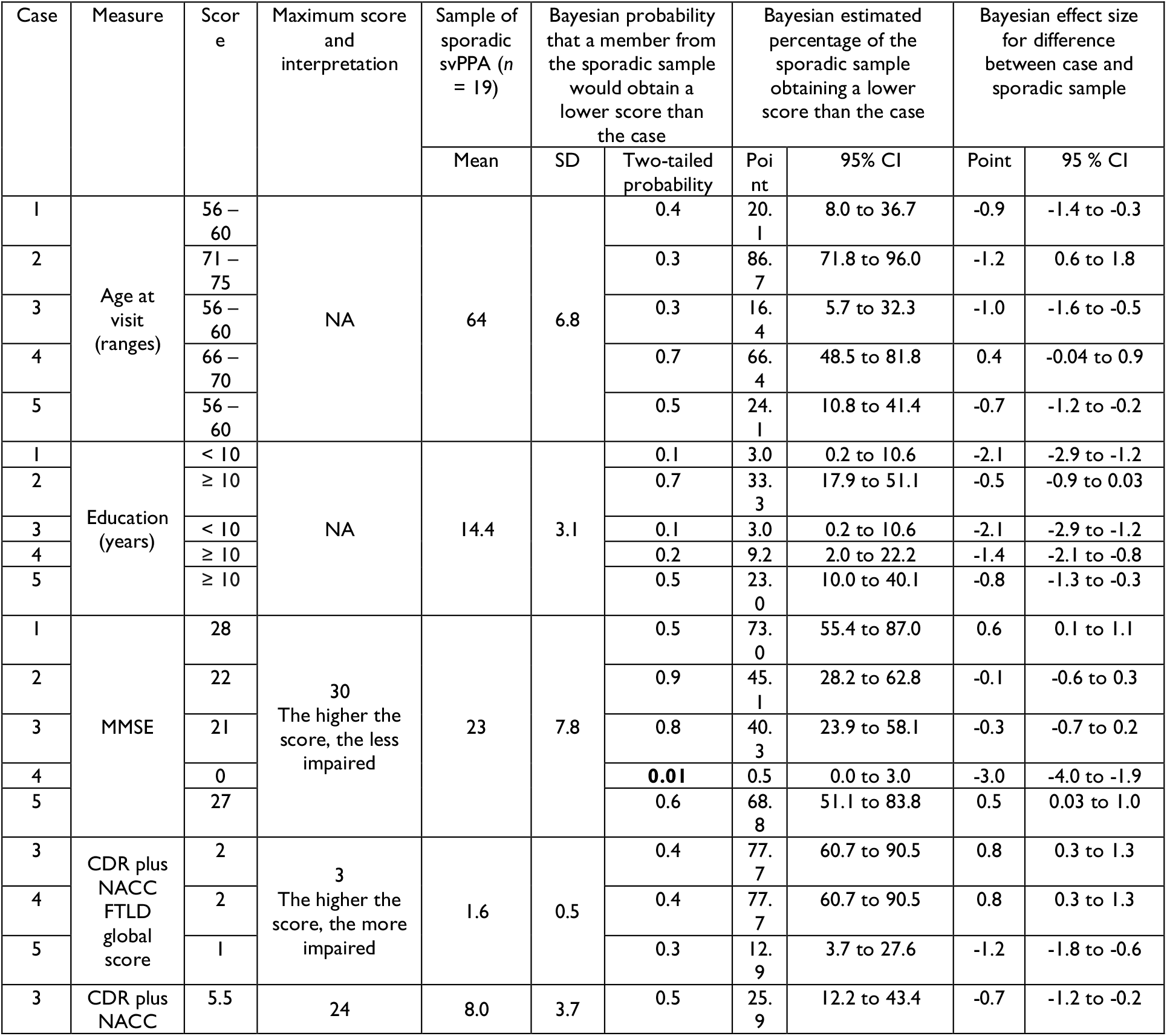
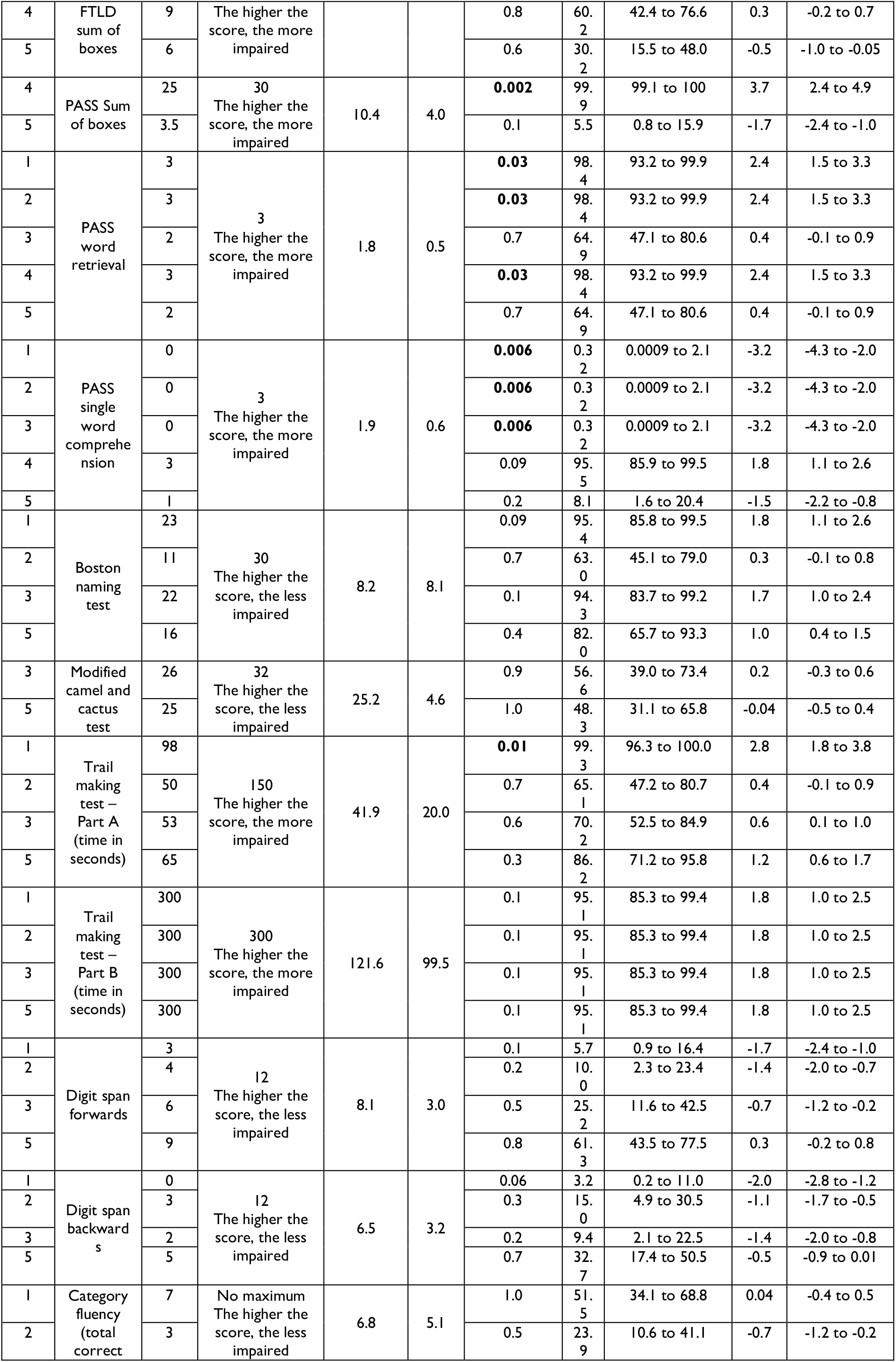
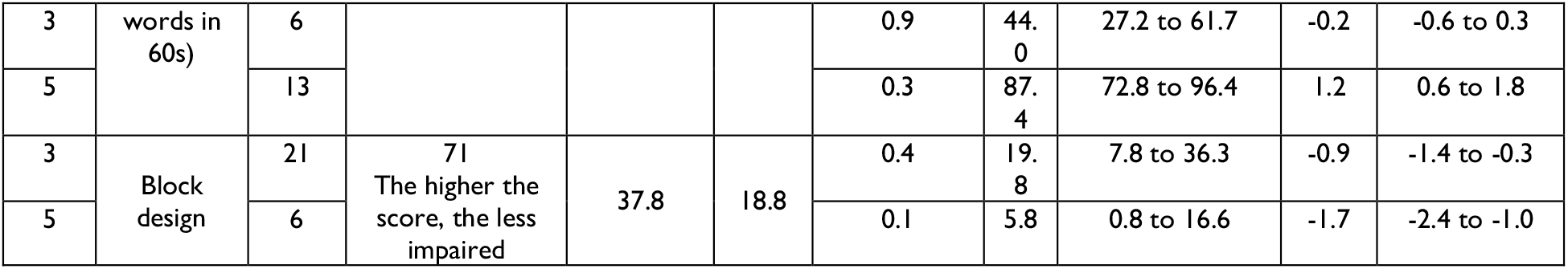
Bayesian point and interval estimates of effect sizes for each GENFI case to svPPA sporadic sample. . GENFI standardised clinical examination includes demographics, clinical symptom data, and neuropsychological assessment

Significant differences between genetic and sporadic cases were notable for the PASS subdomain scores of word retrieval and single word comprehension, two hallmark features of svPPA. Specifically, genetic patients diagnosed as svPPA were rated as being more severely impaired on word retrieval and more preserved on single word comprehension. The Bayesian estimated percentage of the sporadic sample obtaining a lower score than the genetic cases on PASS word retrieval ranged between 65% and 98%. This highlights the unusually high rating on the word retrieval domain (reflecting a greater impairment) for the GENFI cases. In contrast, four out of five cases had a score of 1 or 0 on the PASS single word comprehension domain and the estimated percentage of the sporadic sample obtaining a lower score than this ranged between 0.3% and 8%.

On the other hand, genetic svPPA patients performed better than the sporadic sample on the Boston Naming Test as shown in the high estimated percentage (ranging between 63% and 95%) and effect sizes (ranging between 0.3 and 1.8 with an average of 1.2). Of further note, genetic patients classified as svPPA performed poorly on executive tasks such as the Trail Making Test Parts A and B, as well as Digit Span, both forward and backward, relative to the svPPA sporadic sample. In the large literature on sporadic SD, digit span is typically preserved for a substantial time in SD (see for example Shebani and Patterson^22^ and Coemans *et al*.^23^).

Significant two-tailed probabilities are indicated in bold. Note: Correct interpretation depends on the scoring of each measure. For example, higher values on the PASS denote greater impairment, whereas higher raw scores on standard tests indicate better performance. For Trail Making Test A and B, the total score is the time it took each participant to complete the task. The estimated percentages and effect sizes should be interpreted accordingly. As an example, if the estimated percentage of the sporadic sample obtaining a lower score than the GENFI case on Trail Making Test is high, this means that the sporadic sample would have a lower value for the total time, indicating better performance.

### Systematic review

Table 3 provides a summary of the 20 publications reporting familial/genetic SD/svPPA. The eight publications that included MAPT mutations identified 14 cases, where P301L mutation accounted for the majority (10/14). The average age of onset was 48 with a range of 37 to 69 years and there were more males (9/14) than females (5/14). Semantic deficits were reported to be the first and most prominent symptom for all cases. Details of the presence and stage-of-onset of other symptoms are specified in Table 3. Except two cases, all had structural brain imaging for which anterior temporal lobe (ATL) atrophy was reported. As shown in Table 4, seven cases had a modified Goldman score of 1, delineating an autosomal dominant pattern of inheritance, one had a score of 2, five had a score of 3, and two had a score of 4, where family history was not reported in one and negative in the other. The average modified Goldman score was 2.2.

**Table 3.**
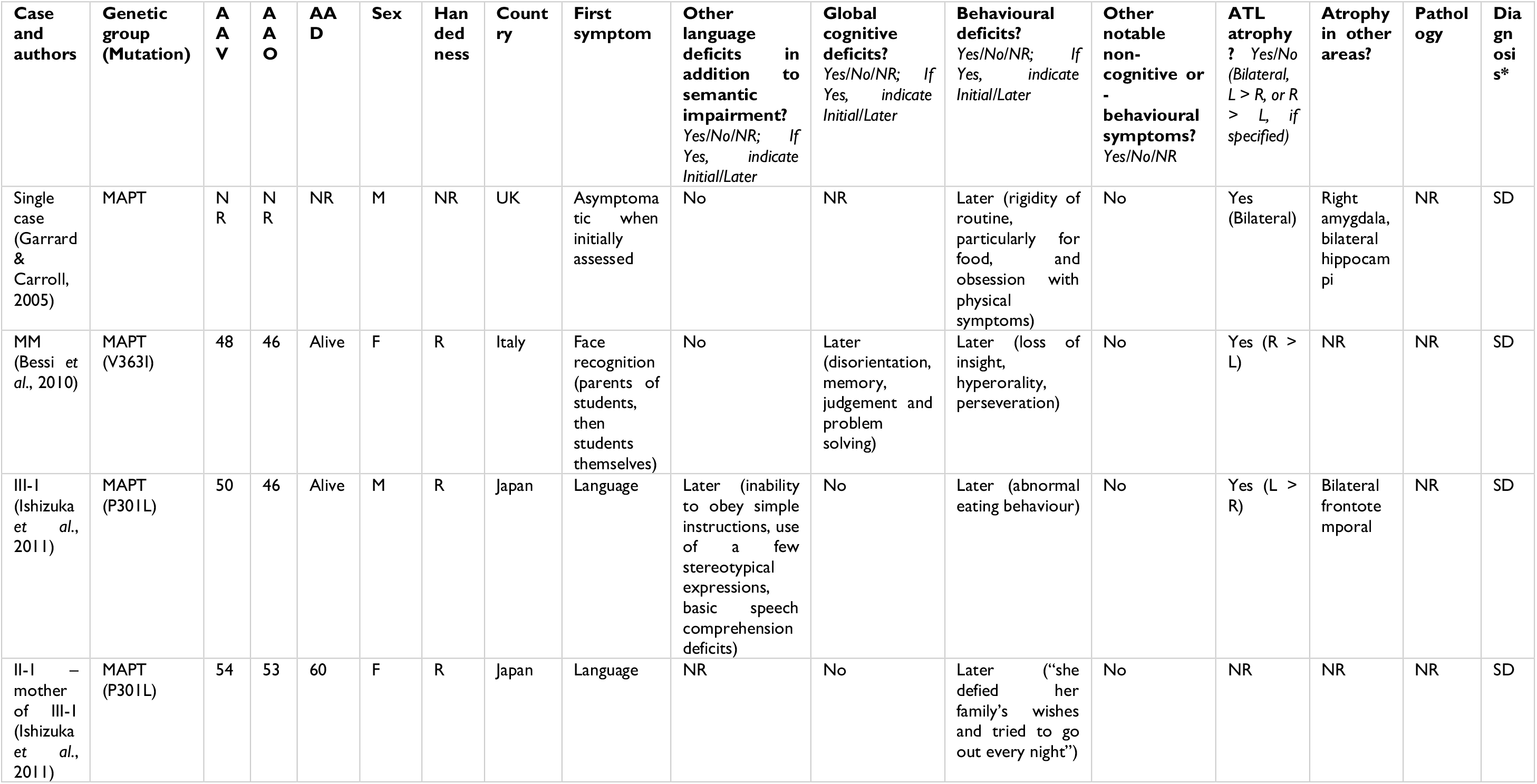
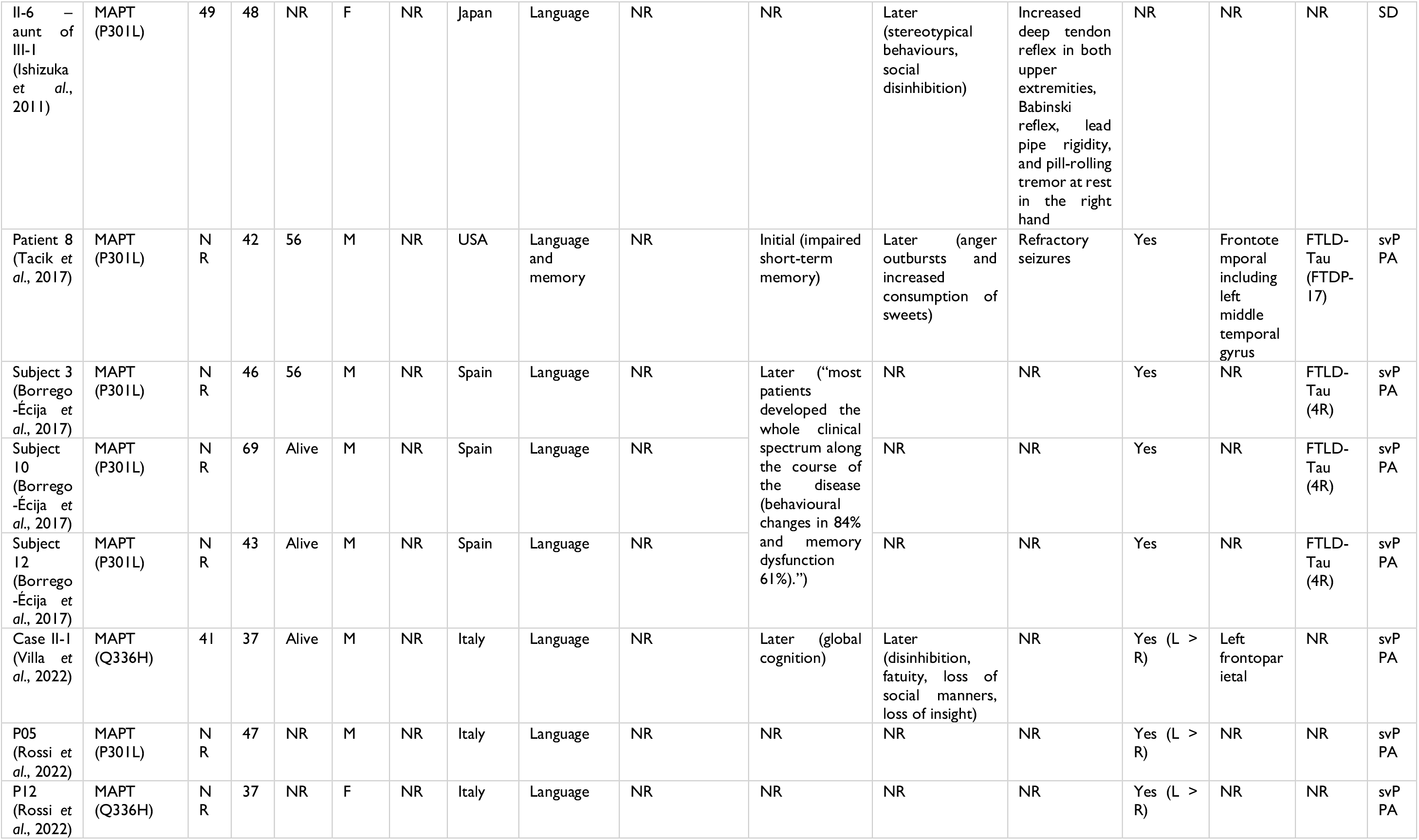
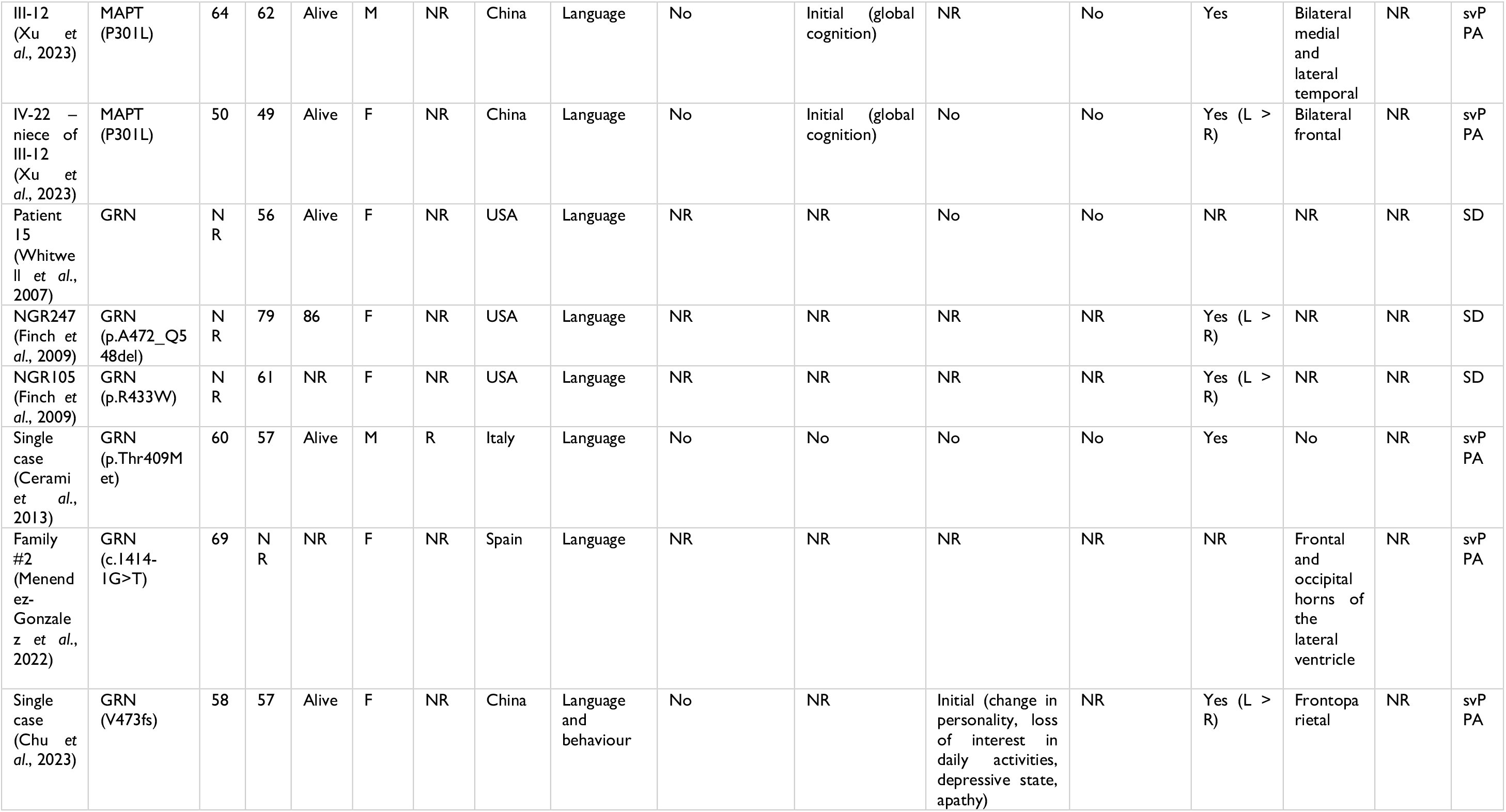
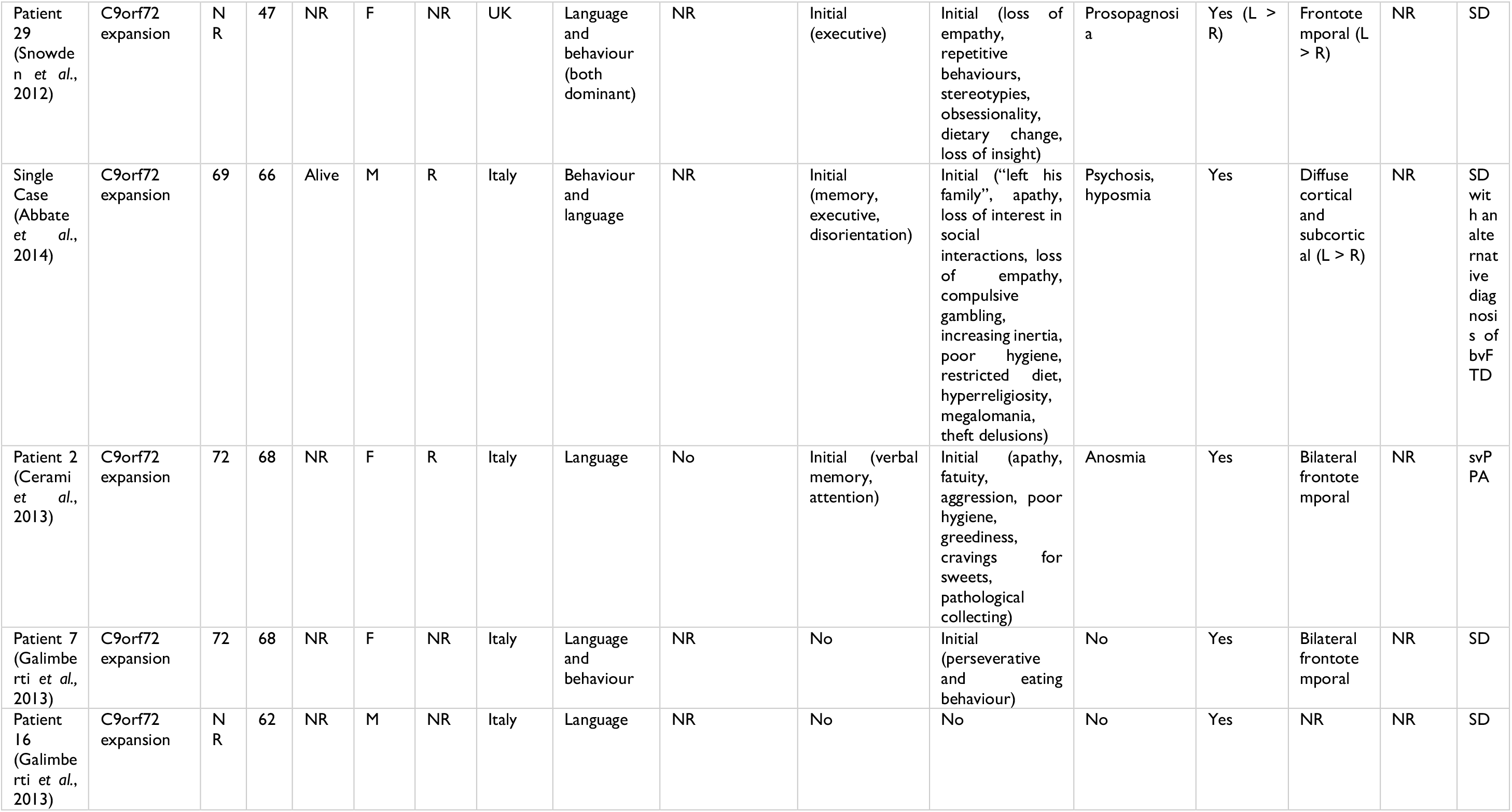
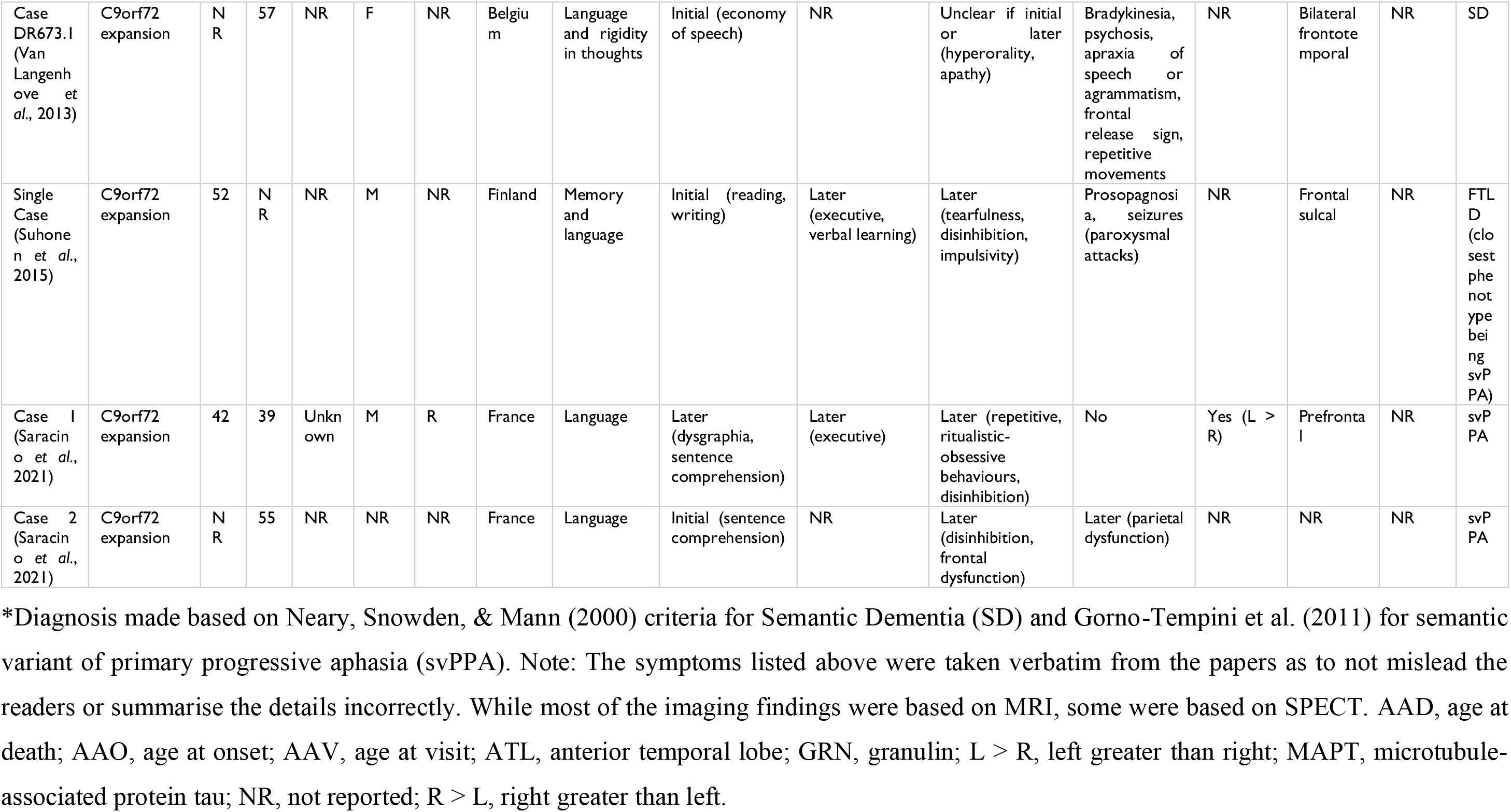
Summary of studies including SD/svPPA cases with MAPT, GRN, or C9orf72 mutations.

**Table 4.** Cases in the published literature with mutations stratified according to family history

| Case and authors | Genetic group (Mutation) | Modified Goldman score | Family details |
| --- | --- | --- | --- |
| Single case (Garrard & Carroll, 2005) | MAPT | 1 | Two brothers with a splice site mutation at exon 10+16 of MAPT with a progressive behavioural syndrome with akinetic-rigid features and anecdotal evidence of early-onset behavioural change among members of previous generation (including proband's father) |
| MM (Bessi et al., 2010) | MAPT (V363I) | 4 | No family history |
| III-1 (Ishizuka et al., 2011) | MAPT (P301L) | 1 | Five affected individuals in two generations including the proband, his mother, two aunts and an uncle (exhibiting 'pure SD') |
| II-1 – mother of III-1 (Ishizuka et al., 2011) | MAPT (P301L) | 1 |  |
| II-6 – aunt of III-1 (Ishizuka et al., 2011) | MAPT (P301L) | 1 |  |
| Patient 8 (Tacik et al., 2017) | MAPT (P301L) | 1 | Father, brother, sister, two paternal aunts, and three cousins with early onset dementia (as shown in Table 1 of Tacik et al., 2017) |
| Subject 3 (Borrego-Écija et al., 2017) | MAPT (P301L) | 4 | Not reported |
| Subject 10 (Borrego-Écija et al., 2017) | MAPT (P301L) | 3 | Two family members, one with bvFTD and another with AD, but relationship status unknown |
| Subject 12 (Borrego-Écija et al., 2017) | MAPT (P301L) | 3 | One family member with bvFTD but relationship status unknown |
| Case II-1 (Villa et al., 2022) | MAPT (Q336H) | 3 | Parent with AD but age of onset unknown |
| P05 (Rossi et al., 2022) | MAPT (P301L) | 3 | Family history is indicated as “positive”, but details not reported (as shown in Table 3 of Rossi et al., 2022) |
| P12 (Rossi et al., 2022) | MAPT (Q336H) | 3 | Family history is indicated as “positive”, but details not reported (as shown in Table 3 of Rossi et al., 2022) |
| III-12 (Xu et al., 2023) | MAPT (P301L) | 1 | Four generations of individuals exhibiting primary semantic impairments with multiple individuals meeting diagnostic criteria for svPPA |
| IV-22 – niece of III-12 (Xu et al., 2023) | MAPT (P301L) | 1 |  |
| Patient 15 (Whitwell et al., 2007) | GRN | 1 | All eight cases including Patient 15 (the only case with SD) listed as having “an autosomal dominant pattern of inheritance” but details not reported |
| NGR247 (Finch et al., 2009) | GRN (p.A472_Q548del) | 4 | No family history |
| NGR105 (Finch et al., 2009) | GRN (p.R433W) | 3 | Family history indicated as “yes” but details not reported (as shown in Table 1) |
| Single case (Cerami et al., 2013) | GRN (p.Thr409Met) | 2 | Mother with psychotic disorder (auditory and visual hallucination) and memory impairments, maternal grandmother with late-onset dementia. (Note: The authors state that the proband had three additional individuals affected over two generations, but the pedigree (Figure 1) does not reflect this) |
| Family #2 (Menendez-Gonzalez et al., 2022) | GRN (c.1414-1G>T) | 4 | No family history |
| Single case (Chu et al., 2023) | GRN (V473fs) | 3 | Mother with dementia at 65 who died at 70 |
| Patient 29 (Snowden et al., 2012) | C9orf72 expansion | 3 | Mother with FTD |
| Single Case (Abbate et al., 2014) | C9orf72 expansion | 4 | No family history |
| Patient 2 (Cerami et al., 2013) | C9orf72 expansion | 4 | No family history |
| Patient 7 (Galimberti et al., 2013) | C9orf72 expansion | 4 | No family history |
| Patient 16 (Galimberti et al., 2013) | C9orf72 expansion | 4 | No family history |
| Case DR673.I (Van Langenhove et al., 2013) | C9orf72 expansion | 1 | Case listed as having “autosomal dominant inheritance”, but details not reported |
| Single Case (Suhonen et al., 2015) | C9orf72 expansion | 4 | No family history |
| Case 1 (Saracino et al., 2021) | C9orf72 expansion | 4 | No family history (Note: Although not FTLD, MND, CBS or PSP, family history of psychosis noted in a cousin) |
| Case 2 (Saracino et al., 2021) | C9orf72 expansion | 4 | Not reported |

The five publications that reported GRN mutation identified six cases. The average age of onset was 63 with a range of 57 to 79 years and the cases were mainly female (5/6). Semantic deficits were initial in all six cases, although one case also had initial behavioural deficits. Later behavioural or cognitive deficits were either absent or not reported. Four cases had imaging available showing ATL atrophy. One case had a modified Goldman score of 1, one had a score of 2, two had a score of 3, and one had a score of 4, where family history was negative. The average modified Goldman score was 2.8.

The seven publications that reported C9orf72 mutation identified nine cases. The average age of onset was 58 with a range of 39 to 68 years and an even split for sex (4/9 male, 4/9 female, one not specified). Concomitant language and behavioural changes were found in three cases, and an additional four cases later developed behavioural deficits including disinhibition, apathy, impulsivity and/or perseverative behaviour. Other notable symptoms included prosopagnosia in two, hyposmia/anosmia in two, psychosis in two and apraxia of speech (AOS) in one. AOS would be a highly atypical feature in SD: apart from the dramatically empty quality of speech, with many or most specific nouns and verbs replaced by more general, high-frequency terms like *thing* or *place* for nouns and *do*, *go* or *come* for verbs, SD patients’ speech production is fairly normal in phonology and syntax. Imaging was not available for three cases; the rest reportedly showed bilateral ATL atrophy. One case, listed as having an autosomal dominant pattern of inheritance although without details regarding affected family members, was assigned a score of 1; one had a score of 3, and the rest had a score of 4, where family history was not reported in only one case. The average modified Goldman score was 3.6.

## Discussion

Semantic dementia (SD)/semantic variant of primary progressive aphasia (svPPA) is typically sporadic and a family history to suggest autosomal dominant genetic aetiology is rare. We explored the genetics of SD/svPPA in two complementary ways. First, we outlined the clinical, cognitive and imaging normality of two individuals self-reported to be monozygotic (MZ) twins of clear cases of SD/svPPA. Second, we reviewed the clinical and cognitive profile of people who had been classified as SD/svPPA with an FTD-associated genetic mutation. The two affected twin cases displayed all of the characteristics of classic SD/svPPA, in imaging (relatively selective bilateral ATL atrophy) and cognition (profound anomia, impaired word and object comprehension, without prominent initial behavioural change). Their healthy twins were normal in imaging and cognition, despite a 15-year classic-SD progression of one affected twin. Our review of five cases in GENFI categorised as svPPA and 20 published cases of genetic SD/svPPA highlighted notable differences in clinical features between reported genetic cases and sporadic SD/svPPA.

Twin studies provide an opportunity to evaluate the contribution of genetic and environmental factors in disease. Both twin pairs grew up together, sharing not only genetics but also principal environmental exposures. Their discordance for SD/svPPA resembles previous MZ studies for other aetiologically complex diseases ranging from type 2 diabetes to neurodegenerative conditions.^9–11,24^ While we cannot definitively prove that A* and B* will never develop SD/svPPA, A* remains normal in cognition and imaging over the 15 years since A’s initial symptoms and 13 since A’s diagnosis. Case A typifies a clear SD clinical course: initial difficulty in language and semantic memory as indicated by severe anomia and loss of object knowledge, with otherwise largely preserved cognition and behaviour. A’s MRI revealed left predominant bilateral anterior temporal lobe atrophy. Only later in progression did problems in behaviour (e.g., fixed routines, clock-watching, hoarding behaviour, loss of insight) emerge, a development often observed in sporadic SD. A’s diagnosis remained unchanged as the salient feature of both the initial presentation and the pattern of decline was profound semantic deterioration. Case B presented with bilateral but predominant right temporal atrophy and semantic degradation, prosopagnosia, and behavioural features characteristic of typical R>L SD.^25^ Assessment of B’s unaffected twin B* revealed no hint of cognitive or imaging abnormality.

SD/svPPA cases with a positive family history in GENFI and the literature have had parents, siblings, or other relatives with dementia, but none had an MZ twin. We examined the strength of the family history of the genetic SD/svPPA cases in our systematic review using a modified Goldman score from 1 (indicating a clear autosomal dominant history of SD) to 4 (no or unknown family history): 31% (9/29) of cases had a Goldman score = 1. This contrasts with 0% reported by Rohrer and colleagues,^3^ but reflects the selection of cases by their positive mutation status. The results of our systematic review were largely driven by the MAPT-associated mutation group, where seven out of 14 cases had a modified Goldman score of 1 with sufficient family details. Ishizuka and colleagues identified a P301L mutation in the MAPT gene in a Japanese family with ‘pure SD’ (i.e., presenting with semantic memory impairment preceding any behavioural changes) where five individuals were affected in two generations including the proband, his mother, two aunts, and an uncle.^26^ Similarly, P301L mutation was identified in a Chinese family with a strong family history in four generations where the proband met strict criteria for svPPA.^27^ Unlike the GRN- and C9orf72-associated mutation groups, both probands had a strong family history of SD/svPPA. In our systematic review, most of the cases with a positive family history did not have family members affected by progressive language disorders and these results suggest that svPPA is a rare phenotype in GRN, C9orf72, and, to a lesser extent, MAPT mutation carriers. Moreover, MAPT mutations have been reported to be more common in Asian countries than Western countries.^4,28,29^ Geographical variability in different mutations related to SD therefore warrants further investigation. Perhaps equally important to understanding the prevalence of familial SD is to investigate the phenotypic variations within the mutation. Some mutations have shown variability in their age at onset,^30,31^ demographics such as sex^32^; and progression with some mutations is reported to be slow.^33–35^

Assessment of the similarities and differences between genetic and sporadic SD/svPPA cases in the literature is impeded by many factors such as (a) differences in assessment tools; (b) lack of detailed case histories, test scores, imaging, and/or pathology; (c) difficulty ascertaining whether all critical domains were tested for a precise diagnosis; and (d) uncertainty regarding the diagnosis in some cases. Most of the cases in the C9orf72-associated mutation group had initial behavioural disturbance, with some cases also exhibiting symptoms highly unusual in sporadic SD such as apraxia of speech, hyposmia, or psychosis. Abbate and colleagues conveyed the challenge of disentangling SD and bvFTD and presented an alternative diagnosis of bvFTD for a patient initially diagnosed with SD with a hexanucleotide repeat expansion in the C9orf72 gene.^36^ Similarly, despite the single case presented by Suhonen *et al*. having a diagnosis of SD, the authors indicated that imaging and other behavioural deficits made ‘pure SD’ unlikely, and that SD was simply the closest phenotype.^33^ This diagnostic problem also extends to other mutation groups and to FTD diagnoses more generally. For instance, because prominent semantic impairment is often associated with behavioural symptoms in MAPT-related FTD, it is reported to be difficult to ascertain a diagnosis of SD versus bvFTD.^37^ These reports highlight two key takeaways. First, when the clinical picture does not conform to a clear clinical entity, diagnostic labels may serve as the nearest approximations or a clinical heuristic. Second, the particular overlap between SD and bvFTD emphasises the view that syndromes associated with FTD exist as a multidimensional spectrum, with many or most patients displaying some of the features characteristic of multiple diagnoses.^38,39^

In contrast to the inherent challenges for the systematic literature review, the GENFI data afforded a direct comparison of the neuropsychological, clinical, and symptom data between five mutation-carriers classified as svPPA *versus* a sporadic sample with the same assessments.^12^ Notable differences between genetic and sporadic cases emerged as follows: (i) genetic patients were rated as being more severely impaired on the PASS word retrieval domain (mean = 2.6) relative to the sporadic sample (mean = 1.8) yet the genetic patients performed better on the Boston Naming Test (mean = 18) than the sporadic sample (mean = 8.2); (ii) three genetic patients were rated as being preserved on the PASS single word comprehension domain: yet impaired word comprehension is one of the two hallmark features required to meet the current diagnostic criteria for svPPA^17^; and (iii) genetic patients performed poorly on executive tasks relative to the svPPA sporadic sample.

The only individual in GENFI bearing a mutation in the GRN gene was Case 4 who showed significantly impaired scores on the MMSE and the PASS sum of boxes compared to the sporadic svPPA sample. The missing neuropsychological data and structural MRI may reflect this individual’s severe global impairment. Case 4’s C253X mutation has only been reported in five cases worldwide with each having a different clinical diagnosis: svPPA, bvFTD, Alzheimer’s disease, dementia with Lewy bodies, and dementia not otherwise specified.^4^ In other words, less is known about the mutation’s phenotypic variability and clinical picture. Case 5’s mutation in the MAPT gene is also distinctive given that this p.P397S variant has been found in five families diagnosed with bvFTD plus significant semantic impairment.^35^ One of these (Family III) included dizygotic twin pairs who were both diagnosed with bvFTD and had a positive family history, with their mother developing dementia at the age of 65 and another sibling showing behavioural dysfunction suggestive of bvFTD. To our knowledge, Case 4 and Case 5 from GENFI are the only ones in the literature bearing these unique mutations with a diagnosis of svPPA.

There were limitations to our study. First is the small set of cases, which is a limitation but also noteworthy as mentioned above. That is, GENFI is a large-scale (*n*>1300), international cohort and it is a striking finding that it includes only five individuals designated as svPPA and with a genetic mutation. A second limitation is that we are only able to present clinical, not pathological, diagnoses. However, clinico-pathological correlations are high in SD/svPPA with TDP-43 Type C as the most likely molecular pathology.^40^ Third, some of the GENFI assessments were missing for a few cases and imaging was available for only three patients. While the missing data preclude a comprehensive comparison between sporadic and genetic cases, we acknowledge that this is a common methodological problem in FTD, where often it is not possible to administer a full formal test battery plus neuroimaging due to the severity of language, behaviour, or other disturbances, or to limited willingness/availability for assessment.

Extensive publications on heritability of traits in the developmental twin literature make a major distinction between DZ and MZ twins, and greater concordance for disease might be expected for MZ twins. The discordance in our twin pairs is therefore consistent with the sporadic nature of SD. The opportunity for longitudinal tracking of MZ twin pairs discordant for SD is rare. Over 30 years in our clinic, we have seen three additional MZ twins with classical SD, whose twin siblings were said to be entirely healthy; but we were not able to assess the unaffected siblings in person. We believe that the detailed follow up and case histories of the two MZ twin pairs reported here may be representative of other MZ twin pairs, and that they highlight both the strongly sporadic nature of SD and current limitations in understanding non-genetic factors that influence SD.

In conclusion, these complementary analyses of twins and cases with a genetic mutation support the essentially sporadic nature of SD/svPPA. There remain gaps in the understanding of environmental and epigenetic influences on sporadic SD and a need for further unbiased genotyping and phenotyping of SD/svPPA and “SD-like” syndromes.

## Data availability

The authors confirm that the derived data supporting the findings of this study are available within the article and its supplementary material.

## Acknowledgements

We thank our patients and their families for supporting this work.

## Supplementary material

**Supplementary Table 1:** Details from the standard GENFI clinical report for the five casess

| **GENFI Case** | **1** | **2** | **3** | **4** | **5** |
| --- | --- | --- | --- | --- | --- |
| **Genetic mutation** | **C9orf72** | **C9orf72** | **C9orf72** | **GRN** | **MAPT** |
| Age at onset (range) | 51 – 55 | 71 – 75 | 56 – 60 | 61 – 65 | 56 – 60 |
| First symptom | Language – decreased fluency | Language – impaired word retrieval | Language – impaired word retrieval | Language – impaired sentence comprehension | Language – impaired word retrieval |
| First symptom 1 | NA | Cognition – memory | Language – decreased fluency | Language – impaired functional communication | Cognition – memory |
| First symptom 2 | NA | Cognition – Impaired judgement problem solving | Cognition – bradyphrenia | Language – impaired speech repetition | Behaviour – apathy |
| Current primary working diagnosis | PPA | PPA | PPA | PPA | PPA |
| Gorno-Tempini (PPA) criteria | svPPA | svPPA | svPPA | svPPA | svPPA |
| Rascovsky (bvFTD) criteria | NA | NA | NA | NA | NA |
| El Escorial (MND/ALS) criteria | NA | NA | NA | NA | NA |
| **1. Behaviour affected?** | Yes | Yes | Yes | No | Yes |
| Disinhibition | Absent | Mild | Absent | NA | Mild |
| Apathy | Moderate | Severe | Absent | NA | Mild |
| Empathy | Severe | Moderate | Absent | NA | Absent |
| OCD | Moderate | Absent | Absent | NA | Absent |
| Appetite | Severe | Absent | Absent | NA | Absent |
| Poor response to social/emotional cues** |  |  | Absent | NA | Absent |
| Inappropriate trusting behaviour** |  |  | Mild | NA | Mild |
| **2. Neuropsychiatric symptom affected?** | Yes | Yes | No | Yes | No |
| Neuropsychiatric** |  |  | NA | Absent | NA |
| Visual hallucinations | Very Mild | Absent | NA | Absent | NA |
| Auditory hallucinations | Absent | Absent | NA | Absent | NA |
| Tactile hallucinations | Absent | Absent | NA | Absent | NA |
| Delusions | Mild | Absent | NA | Absent | NA |
| Depression | Moderate | Moderate | NA | Absent | NA |
| Anxiety | Absent | Mild | NA | Mild | NA |
| Irritability/lability** |  |  | NA | Absent | NA |
| Agitation/aggression** |  |  | NA | Absent | NA |
| Euphoria/elation** |  |  | NA | Absent | NA |
| Aberrant motor behaviour** |  |  | NA | Absent | NA |
| Hypersexuality** |  |  | NA | Absent | NA |
| Hyperreligiosity** |  |  | NA | Absent | NA |
| Impaired sleep** |  |  | NA | Absent | NA |
| Altered sense of humour** |  |  | NA | Absent | NA |
| **3. Language affected (based on PASS ratings)?** | Yes | Yes | Yes | Yes | Yes |
| Impaired articulation | Absent | Absent | Absent | Moderate | Absent |
| Decreased fluency<sup>a</sup> | Moderate | Mild | Mild | Severe | Absent |
| Impaired grammar/syntax | Severe | Mild | Absent | Severe | Absent |
| Impaired word retrieval | Severe | Severe | Moderate | Severe | Moderate |
| Impaired speech repetition | Very mild | Moderate | Absent | Severe | Absent |
| Impaired sentence comprehension | Very mild | Absent | Absent | Severe | Absent |
| Impaired SWC | Absent | Absent | Absent | Severe | Mild |
| Dyslexia | Absent | Moderate | Very mild | Mild | Absent |
| Dysgraphia | Moderate | Absent | Moderate | Mild | Absent |
| Impaired functional communication | Moderate | Moderate | Absent | Severe | Very mild |
| **4. Cognition affected?** | Yes | Yes | Yes | Yes | Yes |
| Memory impairment | Absent | Severe | Moderate | Mild | Mild |
| Orientation** |  |  | Mild | Mild | Very mild |
| Visuospatial impairment | Moderate | Absent | Absent | Mild | Absent |
| Impaired judgment/problem-solving | Mild | Severe | Absent | Mild | Mild |
| Impaired attention | Moderate | Very mild | Absent | Mild | Absent |
| Problems with community affairs |  |  | Absent | Moderate | Very mild |
| Problems at home or with hobbies |  |  | Very mild | Mild | Very mild |
| Impaired personal care |  |  | Absent | Absent | Very mild |
| Person recognition difficulty |  |  | Absent | Absent | Mild |
| Impaired topographical memory |  |  | Absent | Absent | Mild |
| Bradyphrenia |  |  | Mild | Mild | Very mild |
| **5. Motor affected?** | Yes | No | No | Yes | No |
| Motor |  |  | NA | Absent | NA |
| Dysarthria | Absent | NA | NA | Very mild | NA |
| Dysphagia | Absent | NA | NA | Absent | NA |
| Tremor | Absent | NA | NA | Absent | NA |
| Slowness | Mild | NA | NA | Absent | NA |
| Weakness | Absent | NA | NA | Absent | NA |
| Gait disorder | Absent | NA | NA | Absent | NA |
| Falls | Absent | NA | NA | Absent | NA |
| Functional difficulties using hands** |  |  | NA | Absent | NA |
| **6. Autonomic and other symptoms present?** |  |  | No | No | No |
| Autonomic |  |  | NA | NA | NA |
| Impaired blood pressure |  |  | NA | NA | NA |
| Gastrointestinal symptom |  |  | NA | NA | NA |
| Impaired thermoregulation |  |  | NA | NA | NA |
| Urinary symptom |  |  | NA | NA | NA |
| Altered responsiveness to pain |  |  | NA | NA | NA |
| Altered perception of sounds or music |  |  | NA | Absent | NA |
| Altered perception of smell or taste |  |  | NA | Absent | NA |
| Persistent unexplained physical symptom |  |  | NA | Mild | NA |
| Impaired breathing |  |  | NA | Absent | NA |
| Seizures | Absent | Absent | Absent | Absent | Absent |
| Stroke or TIA | Absent | Absent | Absent | Absent | Absent |
| TBI | Absent | Absent | Absent | Absent | Absent |
| Hypertension | Absent | Mild | Absent | Absent | Absent |
| Hypercholesterolaemia | Mild | Mild | Absent | Absent | Absent |
| Diabetes | Absent | Absent | Absent | Absent | Absent |
| Smoking** |  |  | Absent | Absent | Absent |
| Excess alcohol use** |  |  | Absent | Absent | Absent |
| Recreational drug** |  |  | Absent | Absent | Absent |
| Autoimmune disease** |  |  | Absent | Absent | Absent |
| **7. Drug history (currently taking meds)\*\*?** |  |  | No | Yes | No |
| No of drugs** |  |  | NA | 1 | NA |
| EMG | Absent | Absent |  |  |  |
| **8. GENFI examination** |  |  |  |  |  |
| Supranuclear gaze palsy | NA | Absent | Absent | Absent | Absent |
| Impaired eyelid function** |  |  | Absent | Mild | Absent |
| Facial weakness** |  |  | Very mild | Absent | Absent |
| Bulbar palsy | NA | Absent | Absent | Absent | Absent |
| Pseudobulbar palsy | NA | Absent | Absent | Absent | Absent |
| Neck weakness** |  |  | Absent | Absent | Absent |
| Neck rigidity** |  |  | Absent | Absent | Absent |
| Respiratory weakness** |  |  | Absent | Absent | Absent |
| Myoclonus | NA | Absent | Absent | Absent | Absent |
| Rest tremor | NA | Absent | Absent | Absent | Absent |
| Postural tremor | NA | Absent | Very mild (both limbs) | Absent | Absent |
| Dystonia | NA | Absent | Absent | Absent | Absent |
| Chorea** |  |  | Absent | Absent | Absent |
| Bradykinesia | NA | Absent | Absent | Absent | Absent |
| Rigidity | NA | Absent | Absent | Absent | Absent |
| Limb apraxia | NA | Mild | Absent | Absent | Absent |
| Alien limb phenomenon | NA | Absent | Absent | Absent | Absent |
| Cortical sensory loss** |  |  | Absent | Absent | Absent |
| Fasciculations UL | NA | Absent | Absent | Absent | Absent |
| Fasciculations LL | NA | Absent | Absent | Absent | Absent |
| Spasticity UL | NA | Absent | Absent | Absent | Absent |
| Spasticity LL | NA | Absent | Absent | Absent | Absent |
| Weakness UL | NA | Absent | Absent | Absent | Absent |
| Weakness LL | NA | Absent | Absent | Absent | Absent |
| Hyperreflexia UL | NA | Absent | Mild (left and right) | Absent | Absent |
| Hyperreflexia LL | NA | Severe | Mild (left and right) | Absent | Absent |
| Ataxia | NA | Absent | Absent | Absent | Absent |
| Abnormal gait | NA | NA | Absent | Absent | Absent |
| Abnormal gait type | NA | NA | NA | NA | NA |
| Abnormal gait severity | NA | NA | NA | NA | NA |
| Memory and Orientation | 20 | 18 | 10 | NA | 12 |
| Everyday skills | 12 | 12 | 0 | NA | 0 |
| Self care | 7 | 0 | 0 | NA | 0 |
| Abnormal behaviour | 8 | 8 | 0 | NA | 1 |
| Mood | 6 | 11 | 2 | NA | 0 |
| Beliefs | 4 | 0 | 0 | NA | 0 |
| Eating habits | 8 | 0 | 0 | NA | 2 |
| Sleep | 2 | 3 | 0 | NA | 0 |
| Stereotypic and motor behaviours | 5 | 2 | 0 | NA | 2 |
| Motivation | 7 | 13 | 0 | NA | 5 |
| CBI Total | 79 | 67 | 12 | NA | 22 |
| Frontotemporal Dementia Rating Scale (FRS) | 23 | 53 | 81 | NA | 57 |
| ALS FRS-R | 99 | 48 | 99 | 43 | 48 |
| Modified Interpersonal Reactivity Index (mIRI) total** |  |  | 36 | NA | 50 |
| mIRI EC** |  |  | 21 | NA | 19 |
| mIRI PT** |  |  | 15 | NA | 31 |
| Revised Self-Monitoring Scale (RSMS) total** |  |  | 31 | NA | 64 |
| RSMS EX** |  |  | 14 | NA | 29 |
| RSMS SP** |  |  | 17 | NA | 35 |
| FTLD CDR SOB** |  |  | 5,5 | 9 | 6 |
| FTLD CDR Global** |  |  | 2 | 2 | 1 |
| CDR SOB** |  |  | 3,5 | 6 | 4 |
| GENFI CDR SOB** |  |  | 5,5 | 9 | 6 |
| GENFI CDR Global** |  |  | 2 | 2 | 1 |
| Behaviour SOB** |  |  | NA | 0 | 3 |
| PASS SOB** |  |  | NA | 25 | 3,5 |
| MMSE** | 28 | 22 | 21 | 0 | 27 |

<sup>a</sup>Fluency is one of the Progressive Aphasia Severity Scale (PASS) domains defined as the “degree to which speech flows easily or is interrupted by hesitations, fillers, and pauses” and is not synonymous to the test of verbal fluency. Double asterisks\*\* indicate domains that were assessed only in GENFI 2 (cf. GENFI 1).

## Appendix

### GENFI Consortium collaborators and affiliations

- Robert Laforce^1^
- Caroline Graff^2,3^
- Rik Vandenberghe^4,5,6^
- Alexandre de Mendonça^7^
- Pietro Tiraboschi^8^
- Isabel Santana^9,10^
- Alexander Gerhard^11,12,13^
- Johannes Levin^14,15,16^
- Sandro Sorbi^17,18^
- Markus Otto^19^
- Florence Pasquier^20^
- Simon Ducharme^21,22^
- Chris R. Butler^23,24^
- Isabelle Le Ber^25,26,27^
- Elizabeth Finger^28^
- Maria Carmela Tartaglia^29^
- Mario Masellis^30^
- Fermin Moreno^31,32,33^
- Rhian Convery^34^
- Martina Bocchetta^34^
- David Cash^34^
- Sophie Goldsmith^34^
- Kiran Samra^34^
- David L. Thomas^35^
- Thomas Cope^36^
- Maura Malpetti^37^
- Antonella Alberici^38^
- Enrico Premi^39^
- Roberto Gasparotti^40^
- Emanuele Buratti^41^
- Valentina Cantoni^38^
- Andrea Arighi^42^
- Chiara Fenoglio^43^
- Vittoria Borracci^42^
- Maria Serpente^42^
- Tiziana Carandini^42^
- Emanuela Rotondo^42^
- Giacomina Rossi^8^
- Giorgio Giaccone^8^
- Giuseppe Di Fede^8^
- Paola Caroppo^8^
- Sara Prioni^8^
- Veronica Redaelli^8^
- David Tang-Wai^44^
- Ekaterina Rogaeva^29^
- Johanna Krüger^45,46^
- Miguel Castelo-Branco^47^
- Morris Freedman^48^
- Ron Keren^49^
- Sandra Black^30^
- Sara Mitchell^30^
- Christen Shoesmith^50^
- Robart Bartha^51,52^
- Rosa Rademakers^53^
- Jackie Poos^54^
- Janne M. Papma^54^
- Lucia Giannini^54^
- Liset de Boer^54^
- Julie de Houwer^54^
- Rick van Minkelen^55^
- Yolande Pijnenburg^56^
- Benedetta Nacmias^57^
- Camilla Ferrari^57^
- Cristina Polito^58^
- Gemma Lombardi^57^
- Valentina Bessi^57^
- Enrico Fainardi^59^
- Stefano Chiti^59^
- Mattias Nilsson^60^
- Henrik Viklund^61^
- Melissa Taheri Rydell^2,3^
- Vesna Jelic^62,63^
- Linn Öijerstedt^2,3^
- Tobias Langheinrich^64,65^
- Albert Lladó^66^
- Anna Antonell^66^
- Jaume Olives^66^
- Mircea Balasa^66^
- Nuria Bargalló^67^
- Sergi Borrego-Ecija^66^
- Ana Verdelho^68^
- Carolina Maruta^69^
- Tiago Costa-Coelho^7,70,71^
- Gabriel Miltenberger^7^
- Frederico Simões do Couto^72^
- Alazne Gabilondo^31,32,73^
- Ioana Croitoru^73,32^
- Mikel Tainta^73,32^
- Myriam Barandiaran^31,32,73^
- Patricia Alves^73,74^
- Benjamin Bender^75^
- David Mengel^76,77^
- Lisa Graf^76^
- Annick Vogels^78^
- Mathieu Vandenbulcke^79^
- Philip Van Damme^80^
- Rose Bruffaerts^81^
- Koen Poesen^82^
- Pedro Rosa-Neto^83^
- Maxime Montembault^84^
- Raphaella Lara Migliaccio^25,26^
- Ninon Burgos^25,85^
- Daisy Rinaldi^25,26,27^
- Catharina Prix^86^
- Elisabeth Wlasich^86^
- Olivia Wagemann^86^
- Sonja Schönecker^86^
- Alexander Maximilian Bernhardt^86^
- Anna Stockbauer^86^
- Jolina Lombardi^87^
- Sarah Anderl-Straub^87^
- Adeline Rollin^88^
- Gregory Kuchcinski^20^
- Maxime Bertoux^20^
- Thibaud Lebouvier^20^
- Vincent Deramecourt^20^
- João Durães^89^
- Marisa Lima^89^
- Maria João Leitão^90^
- Maria Rosario Almeida^91^
- Miguel Tábuas-Pereira^92^
- Sónia Afonso^93^
- João Lemos^91^

1. Clinique Interdisciplinaire de Mémoire, Département des Sciences Neurologiques, CHU de Québec, and Faculté de Médecine, Université Laval, QC, Canada
2. Department of Neurobiology, Care Sciences and Society; Center for Alzheimer Research, Division of Neurogeriatrics, Bioclinicum, Karolinska Institutet, Solna, Sweden
3. Unit for Hereditary Dementias, Theme inflammation and Aging, Karolinska University Hospital, Solna, Sweden
4. Laboratory for Cognitive Neurology, Department of Neurosciences, KU Leuven, Leuven, Belgium
5. Neurology Service, University Hospitals Leuven, Leuven, Belgium
6. Leuven Brain Institute, KU Leuven, Leuven, Belgium
7. Faculty of Medicine, University of Lisbon, Lisbon, Portugal
8. Fondazione IRCCS Istituto Neurologico Carlo Besta, Milano, Italy
9. University Hospital of Coimbra (HUC), Neurology Service, Faculty of Medicine, University of Coimbra, Coimbra, Portugal
10. Center for Neuroscience and Cell Biology, Faculty of Medicine, University of Coimbra, Coimbra, Portugal
11. Division of Psychology Communication and Human Neuroscience, Wolfson Molecular Imaging Centre, University of Manchester, Manchester, UK
12. Department of Nuclear Medicine, Center for Translational Neuro- and Behavioral Sciences, University Medicine Essen, Essen, Germany
13. Department of Geriatric Medicine, Klinikum Hochsauerland, Arnsberg, Germany
14. Department of Neurology, Ludwig-Maximilians Universität München, Munich, Germany
15. German Center for Neurodegenerative Diseases (DZNE), Munich, Germany
16. Munich Cluster of Systems Neurology (SyNergy), Munich, Germany
17. Department of Neurofarba, University of Florence, Italy
18. IRCCS Fondazione Don Carlo Gnocchi, Florence, Italy
19. Department of Neurology, University of Ulm, Germany
20. Univ Lille, France; Inserm 1172, Lille, France; CHU, CNR-MAJ, Labex Distalz, LiCEND Lille, France
21. Douglas Mental Health University Institute, Department of Psychiatry, McGill University, Montreal, Québec, Canada
22. McConnell Brain Imaging Centre, Montreal Neurological Institute, McGill University, Montreal, Québec, Canada
23. Nuffield Department of Clinical Neurosciences, Medical Sciences Division, University of Oxford, Oxford, UK
24. Department of Brain Sciences, Imperial College London, UK
25. Sorbonne Université, Paris Brain Institute – Institut du Cerveau – ICM, Inserm U1127, CNRS UMR 7225, AP-HP - Hôpital Pitié-Salpêtrière, Paris, France
26. Centre de référence des démences rares ou précoces, IM2A, Département de Neurologie, AP-HP - Hôpital Pitié-Salpêtrière, Paris, France
27. Département de Neurologie, AP-HP - Hôpital Pitié-Salpêtrière, Paris, France
28. Department of Clinical Neurological Sciences, University of Western Ontario, London, ON, Canada
29. Tanz Centre for Research in Neurodegenerative Diseases, University of Toronto, Toronto, ON, Canada
30. Sunnybrook Health Sciences Centre, Sunnybrook Research Institute, University of Toronto, Toronto, Canada
31. Cognitive Disorders Unit, Department of Neurology, Hospital Universitario Donostia, 20014, San Sebastian, Spain
32. Center for Biomedical Research in Neurodegenerative Disease (CIBERNED), Carlos III Health Institute, Madrid, Spain
33. Biogipuzkoa Health Research Institute, Neurosciences Area, Group of Neurodegenerative Diseases, 20014 San Sebastian, Spain
34. Department of Neurodegenerative Disease, Dementia Research Centre, UCL Queen Square Institute of Neurology, London, UK
35. Neuroimaging Analysis Centre, Department of Brain Repair and Rehabilitation, UCL Institute of Neurology, Queen Square, London, UK
36. Cambridge University Hospitals NHS Trust, Cambridge UK
37. Department of Clinical Neurosciences, University of Cambridge, Cambridge, UK
38. Centre for Neurodegenerative Disorders, Department of Clinical and Experimental Sciences, University of Brescia, Brescia, Italy
39. Stroke Unit, ASST Brescia Hospital, Brescia, Italy
40. Neuroradiology Unit, University of Brescia, Brescia, Italy
41. ICGEB Trieste, Italy
42. Fondazione IRCCS Ca’ Granda Ospedale Maggiore Policlinico, Neurodegenerative Diseases Unit, Milan, Italy
43. University of Milan, Centro Dino Ferrari, Milan, Italy
44. The University Health Network, Krembil Research Institute, Toronto, Canada
45. Research Unit of Clinical Medicine, Neurology, University of Oulu, Oulu, Finland
46. Neurocenter, Neurology, Oulu University Hospital, Oulu, Finland
47. Faculty of Medicine, ICNAS, CIBIT, University of Coimbra, Coimbra, Portugal
48. Baycrest Health Sciences, Rotman Research Institute, University of Toronto, Toronto, Canada
49. The University Health Network, Toronto Rehabilitation Institute, Toronto, Canada
50. Department of Clinical Neurological Sciences, University of Western Ontario, London, Ontario, Canada
51. Department of Medical Biophysics, The University of Western Ontario, London, Ontario, Canada
52. Centre for Functional and Metabolic Mapping, Robarts Research Institute, The University of Western Ontario, London, Ontario, Canada
53. Center for Molecular Neurology, University of Antwerp
54. Department of Neurology, Erasmus Medical Center, Rotterdam, Netherlands
55. Department of Clinical Genetics, Erasmus Medical Center, Rotterdam, Netherlands
56. Amsterdam University Medical Centre, Amsterdam VUMC, Amsterdam, Netherlands
57. Department of Neuroscience, Psychology, Drug Research and Child Health, University of Florence, Florence, Italy
58. Department of Biomedical, Experimental and Clinical Sciences “Mario Serio”, Nuclear Medicine Unit, University of Florence, Florence, Italy
59. Neuroradiology Unit, Department of Experimental and Clinical Biomedical Sciences, University of Florence, Florence, Italy
60. Department of Clinical Neuroscience, Karolinska Institutet, Stockholm, Sweden
61. Karolinska University Hospital Huddinge
62. Department of Neurobiology, Care Sciences and Society; Division of Clinical Geriatrics, Karolinska Institutet, Stockholm, Sweden
63. Cognitive clinic, Theme inflammation and Aging, Karolinska University Hospital, Solna, Sweden
64. Division of Neuroscience and Experimental Psychology, Wolfson Molecular Imaging Centre, University of Manchester, Manchester, UK
65. Manchester Centre for Clinical Neurosciences, Department of Neurology, Salford Royal NHS Foundation Trust, Manchester, UK
66. Alzheimer’s disease and Other Cognitive Disorders Unit, Neurology Service, Hospital Clínic, Barcelona, Spain
67. Imaging Diagnostic Center, Hospital Clínic, Barcelona, Spain
68. Department of Neurosciences and Mental Health, Centro Hospitalar Lisboa Norte - Hospital de Santa Maria & Faculty of Medicine, University of Lisbon, Lisbon, Portugal
69. Laboratory of Language Research, Centro de Estudos Egas Moniz, Faculty of Medicine, University of Lisbon, Lisbon, Portugal
70. Institute of Molecular Medicine João Lobo Antunes, University of Lisbon, Lisbon, Portugal
71. Research Institute for Medicines, Faculty of Pharmacy, University of Lisbon, Lisbon, Portugal
72. Faculdade de Medicina, Universidade Católica Portuguesa
73. Instituto de Investigación Sanitaria Biogipuzkoa, Neurosciences Area, Group of Neurodegenerative Diseases, San Sebastian, Spain
74. Department of Educational Psychology and Psychobiology, Faculty of Education, International University of La Rioja, Logroño, Spain
75. Department of Diagnostic and Interventional Neuroradiology, University of Tübingen, Tübingen, Germany
76. Department of Neurodegenerative Diseases, Hertie-Institute for Clinical Brain Research and Center of Neurology, University of Tübingen, Tübingen, Germany
77. Center for Neurodegenerative Diseases (DZNE), Tübingen, Germany
78. Department of Human Genetics, KU Leuven, Leuven, Belgium
79. Geriatric Psychiatry Service, University Hospitals Leuven, Belgium; Neuropsychiatry, Department of Neurosciences, KU Leuven, Leuven, Belgium
80. Neurology Service, University Hospitals Leuven, Belgium; Laboratory for Neurobiology, VIB-KU Leuven Centre for Brain Research, Leuven, Belgium
81. Department of Biomedical Sciences, University of Antwerp, Antwerp, Belgium; Biomedical Research Institute, Hasselt University, 3500 Hasselt, Belgium
82. Laboratory for Molecular Neurobiomarker Research, KU Leuven, Leuven, Belgium
83. Translational Neuroimaging Laboratory, McGill Centre for Studies in Aging, McGill University, Montreal, Québec, Canada
84. Douglas Research Centre, Department of Psychiatry, McGill University, Montreal, Québec, Canada
85. Inria, Aramis project-team, F-75013, Paris, France; Centre pour l’Acquisition et le Traitement des Images, Institut du Cerveau et la Moelle, Paris, France
86. Neurologische Klinik, Ludwig-Maximilians-Universität München, Munich, Germany
87. Department of Neurology, University of Ulm, Ulm, Germany
88. CHU, CNR-MAJ, Labex Distalz, LiCEND Lille, France
89. Neurology Department, Centro Hospitalar e Universitario de Coimbra, Coimbra, Portugal
90. Centre of Neurosciences and Cell Biology, Universidade de Coimbra, Coimbra, Portugal
91. Faculty of Medicine, University of Coimbra, Coimbra, Portugal
92. Neurology Department, Centro Hospitalar e Universitario de Coimbra, Coimbra, Portugal; Faculty of Medicine, University of Coimbra, Coimbra, Portugal
93. Instituto Ciencias Nucleares Aplicadas a Saude, Universidade de Coimbra, Coimbra, Portugal

